# A lay teacher-counsellor delivered brief psychological treatment for moderate to severe depression among older adolescents in senior high schools in Ghana (Y-MIND): a feasibility cluster randomized controlled trial

**DOI:** 10.64898/2026.10.08.26364988

**Authors:** Benedict Weobong, John Hodsoll, Anthony Gerald Akanlu, Kenneth Setorwu Adde, Dzifa A. Attah, Judith Adagnera, Timothy Awine, Simon Bawa, Rebecca Jopling, Bernard Afriyie Owusu, Tarisai Bere, Lucy Owusu, Maria Anyorikeya, Primrose Nyamayaro, Sherifdeen Adams, Fabian Achana, Edith Dambayi, Christopher Ayuure, David J. Heller, Peter Badimak Yaro, Philip B. Adongo, Franklin N. Glozah, Raymond Aborigo, Barbara Barrett, Patrick Smith, Moses Kumwenda, Dixon Chibanda, Melanie Abas, Kimberley Goldsmith, *Y-MIND consortium

## Abstract

**Introduction:** Adolescents experience high rates of depression and anxiety, yet globally 80% of adolescents and almost all in low-and middle-income countries (LMICs) lack access to adolescent-friendly treatments. The African Youth in Mind (Y-MIND) set out to adapt the youth version of the Friendship Bench and assess its feasibility for delivery in Ghana.

**Methods:** In this feasibility cluster randomised controlled trial, we recruited participants aged 15–18 years with probable diagnosis of moderate to severe depression ascertained with the Patient Health Questionnaire (PHQ-9) total score > 9 from six Senior High Schools in Navrongo, Ghana. Schools were randomly allocated (1:2) to enhanced usual care (EUC) alone or EUC combined with Y-MIND, prior to recruitment of participants. Primary outcomes were feasibility (intervention delivery; conducting a fully powered trial). Secondary outcomes were intervention appropriateness, acceptability, and probable depression symptom severity and remission on the PHQ-9.

**Results:** Between July and August 2024, we enrolled and allocated 67 participants (43 [64%] to the EUC plus Y-MIND group and 24 [36%] to the EUC alone group), of whom 64 (95.5%) completed the 5-month primary outcome assessment (41 [95.3%] in the EUC plus Y-MIND group and 23 [95.8%] in the EUC alone group). The feasibility of delivering the Y-MIND intervention from the perspective of adolescents had a mean score of 4.3 (0.5). Therapy quality score was 94.8% (95% CI: 93.0–96.6), while therapy competence was 86.6% (95% CI: 81.2–92.2). We observed initial clinical benefits, including lower severity of depressive symptoms among adolescents in the Y-MIND group as compared to those in the EUC group, with most participants achieving symptomatic remission at the month 5 follow-up.

**Conclusion:** The Y-MIND school-based psychological treatment model demonstrated good acceptability and feasibility, with high recruitment, retention, treatment adherence, and provider competence. While preliminary findings indicate potential improvements in depressive symptoms, the study was not designed to establish effectiveness. These findings justify progression to a fully powered trial to determine clinical effectiveness, cost-effectiveness, and the scalability of the intervention within routine school settings in LMICs. Trial registration: Clinicaltrials.gov (NCT06740084).

## Introduction

Across the globe, one in seven adolescents experience a mental health condition (1), with depression identified as the second most prevalent condition among young people (2). It is particularly concerning because its onset coincides with a critical period of rapid emotional, behavioural, social, and cognitive development (3). The point prevalence of adolescent depression in Africa is 27% with an interquartile range of 20.1% to 31.1% (4). The annual global loss in human capital attributable to depression and anxiety among young people is estimated at US$ 340.2 billion (5).

In Ghana, the treatment gap exceeds 90% (6), reflecting severe shortages of specialist mental health services with fewer than one psychiatrist, psychologist, or other specialist mental health worker per 100,000 population (7,8). Schools therefore represent an important platform for expanding access to evidence-based psychological interventions (9,10), which are recommended as first-line treatments for adolescent depression and have consistently been shown to reduce depressive and anxiety symptoms (11–13).

Although psychological treatments are effective for adolescent depression, most evidence comes from specialist services in high-income countries (13). There is comparatively little evidence regarding culturally adapted structured psychological interventions delivered by non-specialist providers in schools in sub-Saharan Africa (14,15). Furthermore, we found little evidence that school-based psychological interventions for adolescent depression in sub-Saharan Africa have undergone randomized feasibility evaluation of their acceptability and feasibility before progressing to fully powered effectiveness trials.

In Ghana, the Free Senior High School (FSHS) programme has substantially expanded access to secondary education (16,17), while the School Health Education Programme (SHEP) provides the national framework for delivering school health services (18). Of note, guidance and counselling services are well-established in Ghana’s Senior High Schools. However, despite routinely supporting students with emotional distress, suicidal ideation, self-harm, behavioural difficulties, and family problems, Guidance and Counselling Coordinators (GCCs) currently rely largely on non-standardized counselling approaches and have limited access to structured, evidence-based psychological treatments for depression (19). Together, these initiatives create an important opportunity to integrate evidence-based mental health interventions into routine school systems, although implementation of school mental health services remains limited. To address this gap, we developed Y-MIND, a culturally adapted six-session psychological intervention integrating psychoeducation, behavioural activation and problem-solving therapy for adolescents with depression in Ghana. The intervention was developed through a systematic process involving evidence synthesis, stakeholder engagement, and contextual adaptation (20,21). This article reports findings from a trial designed to assess the feasibility and acceptability of the Y-MIND intervention, as well as the study procedures and trial design, to inform a future fully powered randomized efficacy trial.

## Methods

Details of the methodology are described in a published protocol (7); a summary is presented below.

### Study design and participants

This was a parallel, two-arm (Y-MIND plus Enhanced Usual Care versus Enhanced Usual Care) single blinded cluster randomised controlled feasibility trial (RCT) in six Senior High Schools (SHS) across the Kassena-Nankana Municipality and Kassena-Nankana West District, both located in Ghana’s Upper East Region. These districts are largely rural and agrarian, with Kassem and Nankani as the predominant local languages. The Navrongo Health Research Centre (NHRC), which maintains a demographic surveillance system for the area, provided logistical and technical support for recruitment, data collection, and participant tracking. Participants were aged 15-18 years, had a probable diagnosis of moderate to severe depression ascertained with the Patient Health Questionnaire (PHQ-9) total score > 9, and met other *apriori* inclusion and exclusion criteria (7). Our recruitment strategy entailed three key steps: 1. whole-school health education sessions and introduction of the Y-MIND project in the six schools; 2. brief screening using the Patient Health Questionnaire-2 (PHQ-2) (22) – a subset of the PHQ-9 to optimize yield of clinical eligibility for the trial; and 3. participants who scored ≥3 on the PHQ-2 underwent further assessment using the full PHQ-9. Those meeting criteria for probable moderate-to-severe depression were invited to participate in the trial. The PHQ-9 has been previously validated in Ghana (23,24). Participants who gave informed consent (assent for those under 18 years) were interviewed by trained and experienced research assistants to collect data on socio-demographic factors, and school attendance and attainment. Economic outcomes were assessed using the Child and Adolescent Service Use Schedule, EQ-5D-5L, resource use questionnaire, and productivity lost questionnaire, which will be published separately in a forthcoming publication by Owusu et al. Probable anxiety was measured using the 7-item Generalized Anxiety Disorders scale (GAD-7), and the Alcohol Use Disorders Identification Test (AUDIT) was used to assess alcohol use. Data on treatment expectations were planned but were inadvertently omitted during the questionnaire programming process. All surveys were recorded and a subset randomly selected for review by a supervisor for quality assurance. Recruitment was conducted between July and August 2024.

### Sample size estimation

The original sample size for this pilot trial was 60 adolescents across 6 schools (10 in each school). In a school setting, this number of adolescents was deemed feasible to recruit and would provide sufficient data on feasibility, acceptability, and appropriateness of the intervention. Estimates of the standard deviation of Feasibility of Intervention Measure (FIM) (scored as a mean across the items in the FIM scale) from two unpublished studies are 0.34 and 0.59. 60 participants allowed us to estimate the mean FIM score to within a 95% confidence level of between approximately ± 0.09 and ± 0.15 using these two standard deviation estimates.

### Randomization

Six schools were purposively selected out of a possible 13 schools in the study area based on size and gender composition. The randomisation list for the six schools was created by an independent statistician at King’s College London (KCL) (JH) in a 2:1 ratio to either the Y-MIND intervention and enhanced usual care (EUC) (4 schools) or EUC arm only (2 schools). This was then sent to a member of the Ghana team (TA), who applied the allocations to the schools, prior to recruitment. A 2:1 allocation ratio was selected because the study prioritized intervention feasibility over comparative effectiveness. Assigning more schools to the Y-MIND intervention arm increased opportunities to examine delivery across varied school contexts and to assess fidelity, acceptability, provider competence, attendance, and implementation challenges. The control schools primarily allowed testing of recruitment, retention, data-collection, and other comparison-group procedures.

### Interventions

Schools were allocated at random to either of two intervention arms:

#### Enhanced Usual Care (EUC)

Usual care in Ghanaian schools consists primarily of guidance and counselling services delivered by Guidance and Counselling Coordinators (GCCs). While these services provide important emotional, academic, and psychosocial support, GCCs currently lack access to standardized, evidence-based psychological interventions for adolescent depression, including screening tools. The usual care and treatment provided by the GCCs was enhanced by providing a brief training to recognise and manage poor mental health in adolescents based on the WHO School Health Services Manual and mhGAP Intervention Guides (25,26). This EUC is the most intensive model for school-based mental health care and primary care for mental health and psychosocial conditions that may be envisaged in the foreseeable future in Ghana or other LMICs and is currently recommended by the WHO’s mhGAP. Teachers were sensitised on symptoms recognition, basic mental health principles, and referral procedures. Participants identified as having probable moderate to severe depressive symptoms following the initial screening received a feedback letter to their GCC indicating their PHQ-9 score and guidance on seeking additional support. A detailed description of the EUC component is provided in the published trial protocol paper (7).

#### African Youth in Mind (Y-MIND)

Participants received EUC plus Y-MIND, a manualized Psychological Treatment based on Psychoeducation, Behavioural Activation (BA) and Problem-Solving Therapy (PST) that includes the following strategies: therapeutic alliance, psychoeducation, collaborative goal setting and homework, activity monitoring and scheduling, structured problem solving, positive reinforcement and social support strengthening. Y-MIND was adapted from the youth version of the Friendship Bench (YouFB), a problem-solving therapy-based intervention that has demonstrated promising effectiveness among young people in Zimbabwe (27,28). Based on extensive formative and pilot research, we made several adaptations to strengthen the theoretical orientation and enhance the contextual acceptability and feasibility of the treatment, notably: integrating Behavioral Activation Therapy (20), and introducing cultural norms of distress, values (i.e. family, religion, respect, responsibility), idioms, metaphors and activities (i.e., drumming, storytelling) related to healing and recovery. Y-MIND’s theory is that when adolescents develop emotional and cognitive insight into their mental health challenges and how these can contribute to depression (through psychoeducation), they begin to reduce self-blame and understand their experiences better. By re-engaging reward pathways and countering withdrawal through behavioral activation, they rebuild motivation and a sense of connection. Through problem-solving therapy, they strengthen perceived self-control, problem-coping confidence, and emotional regulation skills. Together, these processes lead to measurable improvements in mood, self-efficacy, and daily functioning, while supporting relapse prevention and sustained well-being. Y-MIND was delivered in an individual format and involved six weekly sessions. Sessions were conducted face-to-face over three phases, at a designated counselling room in the school premises. The beginning phase, comprising 1 session focused on initial engagement and orientation to treatment, its theory, and psychoeducation. The middle phase consisted of a multi-session series (sessions 2-5) concerned with skill acquisition (BA and PS) and active change. The ending phase, consisting of 1 session, centred on reviewing triggers, consolidating skills and future planning to prevent relapse. Each participant was offered all six sessions. A detailed description of the approach and content of each session is provided elsewhere (20). Adolescents who missed three consecutive sessions were considered to have dropped out of treatment. Adolescents who dropped out of treatment did not meet treatment completion criteria and were categorised as unplanned discharge. Adolescents who did not respond (i.e. had consecutive high scores on the PHQ-9 over two sessions and/or reported suicidal behaviour) were referred for specialist care. Supervision was provided weekly by the study psychologists.

#### Interventionists

Like the EUC arm, Y-MIND was delivered by Guidance and Counselling Coordinators (GCCs) in the intervention schools, above 18 years of age, with no professional mental health training and had been officially assigned by their respective school to play a counselling role. The GCCs underwent a one-week intensive training program and a six-week Y-MIND internship, supervised by two licensed clinical psychologists. Training was conducted by two local specialists and two international experts in Behavioural Activation and Problem-Solving Therapy. Supervision was conducted weekly, in two modes: face-to-face at the respective schools, and virtually as a group. Each intervention school had two GCCs. Intervention fidelity and GCC clinical competence were assessed using an adapted version of the Problem-Solving Therapy Fidelity (PROOF) scale (29). A shortened PROOF fidelity and competency checklist (Y-MIND Fidelity and Competency Checklist – Y-MIND FCC) was generated. Fidelity items were scored as 0 (session component not completed) or 1 (session component completed). Competency items were scored as 0 (competency not demonstrated) or 1 (competency demonstrated). The two licensed psychologists piloted the measure and assessment process using randomly selected audio recorded sessions to establish inter-rater consistency, with discrepancies resolved by consensus following independent review. The finalized procedures were then applied to 16 randomly selected 20% of audio recorded sessions. The Y-MIND intervention was delivered between July 2024 and September 2024.

### Outcomes

Primary and secondary outcome data were collected within 4 weeks of the 5 months post-baseline timepoint between December 2024 and January 2025. The primary outcome for feasibility was: 1) the feasibility of the intervention from the perspective of the adolescents (i.e. the extent to which the Y-MIND intervention can be successfully used or carried out within the study setting) assessed by the 4-item Feasibility of Intervention Measure (FIM) (30) defined as the median (IQR) FIM score (completely disagree=1; disagree=2; neither agree nor disagree=3; agree=4; completely agree=5); 2) feasibility of conducting a fully powered trial based on the following parameters: the number and proportion of adolescents screened, number and proportion completing the eligibility assessment, number and proportion eligible, proportion who consented, number of weeks to complete the intervention, and proportion of those who consented and enrolled with clinical and self-report assessments at 5 months follow up. Secondary feasibility outcome was the feasibility of the intervention from the perspective of the GCCs. Other implementation outcomes included: appropriateness of Y-MIND (i.e. extent to which the perceived fit, relevance, or compatibility of Y-MIND for treating adolescents with depression in the study setting) assessed by the 4-item Intervention Appropriateness Measure (IAM) defined as the median (IQR) IAM score (similar response options as the FIM) (30); acceptability of Y-MIND (i.e. the perception among implementation stakeholders that Y-MIND meets their approval, is appealing, liked and welcome) assessed by the 4-item Acceptability of Intervention Measure (AIM) defined as the median (IQR) AIM score (similar response options as the FIM) (30) administered to the adolescents and the GCCs.

The primary clinical outcome was depression severity assessed by the 9-item Patient Health Questionnaire PHQ-9). Secondary clinical outcomes were: remission from probable depression defined as PHQ□9 < 10 with ≥5□point reduction from baseline; severity of probable anxiety assessed by GAD-7; severity of alcohol use disorders assessed by (AUDIT). Academic performance defined as mean academic scores in Mathematics, Science, and English; resource use and quality of life estimated from the Child and Adolescent Service Use Schedule, EQ-5D-5L, resource use questionnaire, and productivity lost questionnaire.

### Masking

The independent evaluators who conducted the outcome assessments and had no contact with the schools or other team members were blinded to allocation.

### Statistical methods

The statistical analysis followed a pre-registered Statistical Analysis Plan (SAP) and reporting is aligned to the CONSORT 2010 and the extended guidelines on randomized controlled trials (RCTs) of social and psychological interventions, and pilot/feasibility trials. Analyses followed an intention-to-treat (ITT) approach, including all schools (and consequently participants) in their randomized groups regardless of treatment adherence or discontinuation. School and participant baseline characteristics as well as primary and secondary feasibility measures were summarized with frequencies and percentages or means and SDs or median IQRs, as appropriate.

For the primary clinical outcome, the planned analytic approach was a linear mixed-effects regression model to estimate the individual-level mean group difference in PHQ-9 scores at 5 months, accounting for the hierarchical structure (pupils nested within schools) via a random intercept for school. However, due to model non-convergence resulting from the small number of clusters (n=6), we instead fitted a linear regression model with cluster-robust standard errors. This model estimated the treatment effect adjusted for baseline PHQ-9 scores. Intervention effects are reported as adjusted mean differences (AMDs) with 95% confidence intervals (CI).

As this is a feasibility study, no formal inference tests (p-values) are presented. We report Cohen’s d derived by dividing the AMD by the pooled baseline standard deviation of the outcome, with 95% CI generated in the same way. For binary outcomes, intervention effects are reported as odds ratios (ORs) estimated from mixed-effects logistic regression. No data were imputed (complete case analysis); item-level missing data were pro-rated if <20% of items were missing. Two sensitivity analyses were undertaken: (1) adjusting for age and gender to account for potential imbalances from cluster randomization, and (2) utilizing bootstrap confidence intervals as a robustness check for the small sample size. Analyses were performed using Stata version 18 (Stata Corp, College Station, TX).

A real-world adaptation costing of Y-MIND was conducted from an intersectoral government perspective, reflecting the collaborative resource-sharing arrangements between the education and health sectors under Ghana’s School Health Education Programme (SHEP). The analysis aimed to estimate the first-year financial requirements for integrating Y-MIND into routine school health services across six senior high schools while leveraging existing infrastructure, personnel, and capacity-building systems. We estimated implementation costs using the Cost of Implementing New Strategies (COINS) framework (31) and an activity-based micro-costing approach (32) to identify, quantify, and value resources used during the pre-implementation and implementation phases. We obtained cost data from Y-MIND financial records, procurement documents, attendance registers, government salary scales, and prevailing market prices. We excluded research-specific costs to generate policy-relevant estimates.

### Ethical considerations

Ethical approval for implementation was obtained from King’s College London Research Ethics Committee (RESCM-23/24-38948), Ghana Health Services Research Ethics Committee (GHS-ERC:015/06/24), and the Navrongo Health Research Centre Research Institutional Review Board (NHRCIRB480). Oversight was provided by a Trial Management Group and an independent Data Safety and Monitoring Board. In line with local ethical standards, participants received small tokens including Y-MIND branded notebooks, refreshments, and bars of soap during assessments and sessions to acknowledge their time and effort. The study protocol was registered at Clinicaltrials.gov (NCT06740084).

## Results

### Trial conduct and baseline characteristics

From July to August 2024, 290 adolescents from the six randomised schools were pre-screened with the PHQ-2 to optimize yield of clinical eligibility. Of these, 108 were identified with initial symptoms of depression and were subsequently assessed further using the complete PHQ-9 tool. Of the 67 participants who met the eligibility criteria, 24 were enrolled in the control arm and 43 in the intervention arm (fig 1). There were no refusals at enrolment. Baseline characteristics as shown in Table 1 were similar by arm, except for potential imbalances in gender, source of educational funding, type of dwelling, source of water and food insecurity.

**Fig 1.**
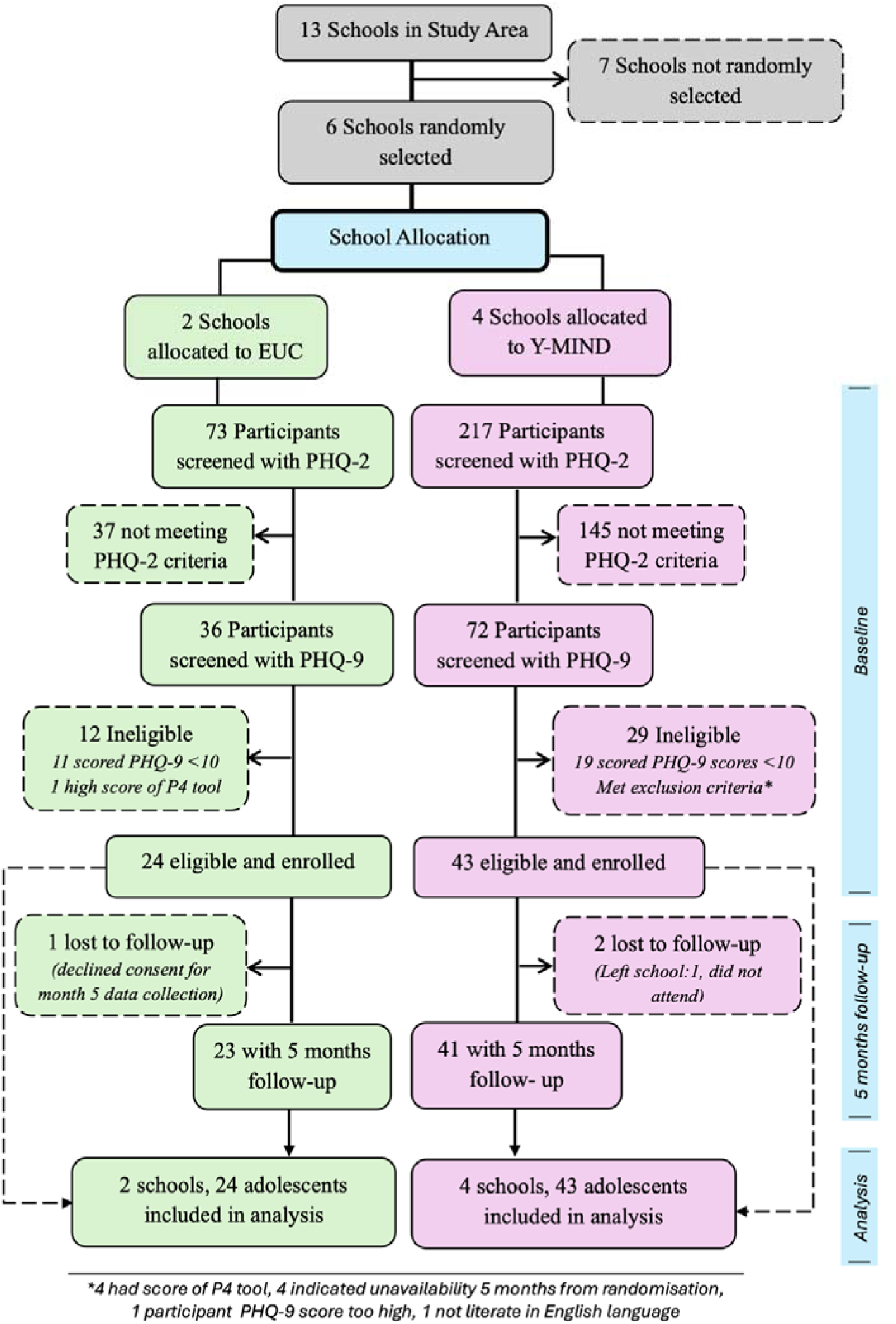
illustrates the trial profile.

**Table 1:** Baseline characteristics (N = 67).

| Variables | Y-MIND<br>(N=43) | EUC<br>(N=24) | Overall<br>(N=67) |
| --- | --- | --- | --- |
| **Age** |  |  |  |
| Mean (SD) | 16.9 (0.8) | 16.8 (0.9) | 16.8 (0.8) |
| **Gender, n(%)** |  |  |  |
| Male | 6 (14.0) | 13 (54.2) | 19 (28.4) |
| Female | 37 (86.0) | 11 (45.8) | 48 (71.6) |
| **Level of Study, n(%)** |  |  |  |
| SHS 1 | 25 (58.1) | 13 (54.2) | 38 (56.7) |
| SHS 2 | 18 (41.9) | 11 (45.8) | 29 (43.3) |
| **School Residential Status, n(%)** |  |  |  |
| Boarding | 42 (97.7) | 24 (100.0) | 66 (98.5) |
| Day | 1 (2.3) | 0 (0.0) | 1 (1.5) |
| **Main Source of Educational Funding, n(%)** |  |  |  |
| Own parents | 29 (67.4) | 13 (54.2) | 42 (62.7) |
| Other guardian | 8 (18.6) | 8 (33.3) | 16 (23.9) |
| Other | 6 (14.0) | 3 (12.5) | 9 (13.4) |
| **Marital status of parents, n(%)** |  |  |  |
| Married and living together | 20 (46.5) | 10 (41.7) | 30 (44.8) |
| Separated | 8 (18.6) | 3 (12.5) | 11 (16.4) |
| Divorced | 4 (9.3) | 4 (16.7) | 8 (11.9) |
| Widow/Widowed | 9 (20.9) | 6 (25.0) | 15 (22.4) |
| Not Applicable | 2 (4.7) | 1 (4.2) | 3 (4.5) |
| **Are both of your parents alive? n(%)** |  |  |  |
| Both Parents Alive | 32 (74.4) | 17 (70.8) | 49 (73.1) |
| Only Father Alive | 3 (7.0) | 2 (8.3) | 5 (7.5) |
| Only Mother Alive | 6 (14.0) | 4 (16.7) | 10 (14.9) |
| Both parents deceased | 2 (4.7) | 1 (4.2) | 3 (4.5) |

1. Note: Standard deviation (SD); Interquartile Range (IQR)

### Feasibility of implementing Y-MIND

The feasibility of delivering the Y-MIND intervention from the perspective adolescents had an average FIM mean score of 4.3 (0.5) (fig 2). The corresponding median (IQR) rating was 4.3 (4.0–4.8). From the perspective of GCCs, the FIM mean score was 4.1 (0.3), and the corresponding median rating was 4.0 (3.9,4.3). Fig. 2 provides summary results for other implementation outcomes – acceptability and appropriateness based on perspectives of adolescents and GCCs.

**Figure 2:**
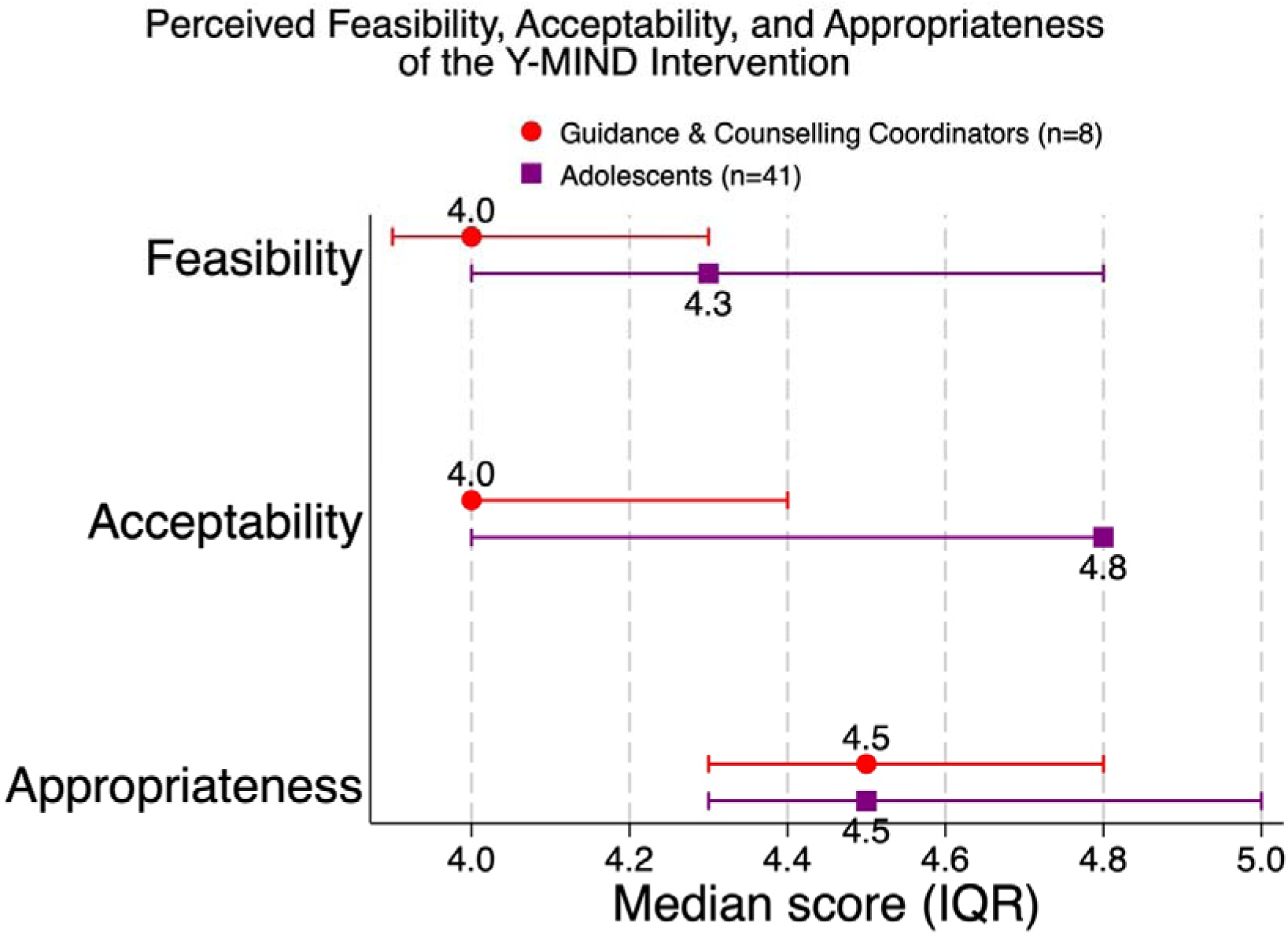
Intervention implementation outcomes

### Therapist competence and therapy quality

A total of eight Guidance and Counselling Coordinators (teachers) were recruited to implement the Y-MIND intervention. Of these, 5 (62.5%) of them were bachelor’s degree holders, and the remaining 3 (37.3%) were master’s degree holders but with no prior training in psychological treatments. The mean therapy quality score reflecting the standard of delivering the Y-MIND intervention by GCCs was 94.8% (95% CI: 93.0–96.6), while their capacity to deliver Y-MIND (competence) was 86.6% (95% CI: 81.2–92.2), indicating a high level of adherence and proficiency to the Y-MIND intervention.

### Feasibility of conducting a fully powered trial

#### Screening and eligibility

A total of 290 adolescents were screened using the PHQ-2 tool. Screening was voluntary and restricted to students in Forms 1 and 2 aged 15-18 years, with eligibility confirmed at the point of screening. Out of the 290, 108 (37.2%) screened positive on PHQ-2 and proceeded to eligibility assessment using the PHQ-9 tool. Of the 108 adolescents assessed for eligibility, 67 (62.0%) met the eligibility criteria for both arms, consented and were enrolled into the study. Thus, we met and exceeded our target sample size of 60 adolescents.

#### Outcome follow-up

Out of the 67 participants enrolled, 64 (95.5%) completed the five-month follow-up assessment, including 23/24 (95.8%) in the control arm and 41/43 (95.3%) in the intervention arm, corresponding to an overall attrition rate of 4.5% (n = 3); they tended to be all males with a mean age of 17 years.

#### Intervention uptake and duration of delivery

Among participants in the intervention arm, 40 out of 43 (93.0%) completed all six sessions, while 3 (7.0%) did not complete the intervention. Among those who completed all sessions, the median time to completion was 6 weeks (IQR: 6-6 weeks), indicating high adherence to the intervention schedule with minimal variability in completion time. The mean session duration was 59.6 min (95% CI 57.1–62.1).

### Clinical outcomes

Results are summarized in fig 2. The mean endpoint PHQ-9 scores was 4.6 (SD 3.6) among adolescents in the Y-MIND arm and 9.9 (SD 5.4) among adolescents in the EUC arm giving a mean group difference of −5.12 points (95% CI: −7.17 to −3.07). Adjustment for age and gender yielded a similar estimate. We also observed that the Y-MIND intervention had a moderate effect in reducing probable anxiety among students as compared to EUC. Specifically, the intervention arm showed a mean difference in GAD-7 score of –3.00 points lower/better as compared to EUC. Participants in the Y-MIND arm also had a higher odds of remission from probable depression than those in the EUC arm with a prevalence difference of 49.4% (95% CI: 27.3 to 71.5).

### Other outcomes

We also explored descriptively the effect of Y-MIND on academic performance in the recent academic year. By 5 months, adolescents in the Y-MIND arm (with 25.6% missing data) showed higher performance in mean scores in Mathematics (52.0 [SD 15.5] vs. 45.0 [SD 27.5]), Science (59.4 [16.1] vs. 43.4 [26.9]), and English (57.1 [14.3] vs. 51.6 [23.9]) compared to those in the EUC arm.

**Fig 3:**
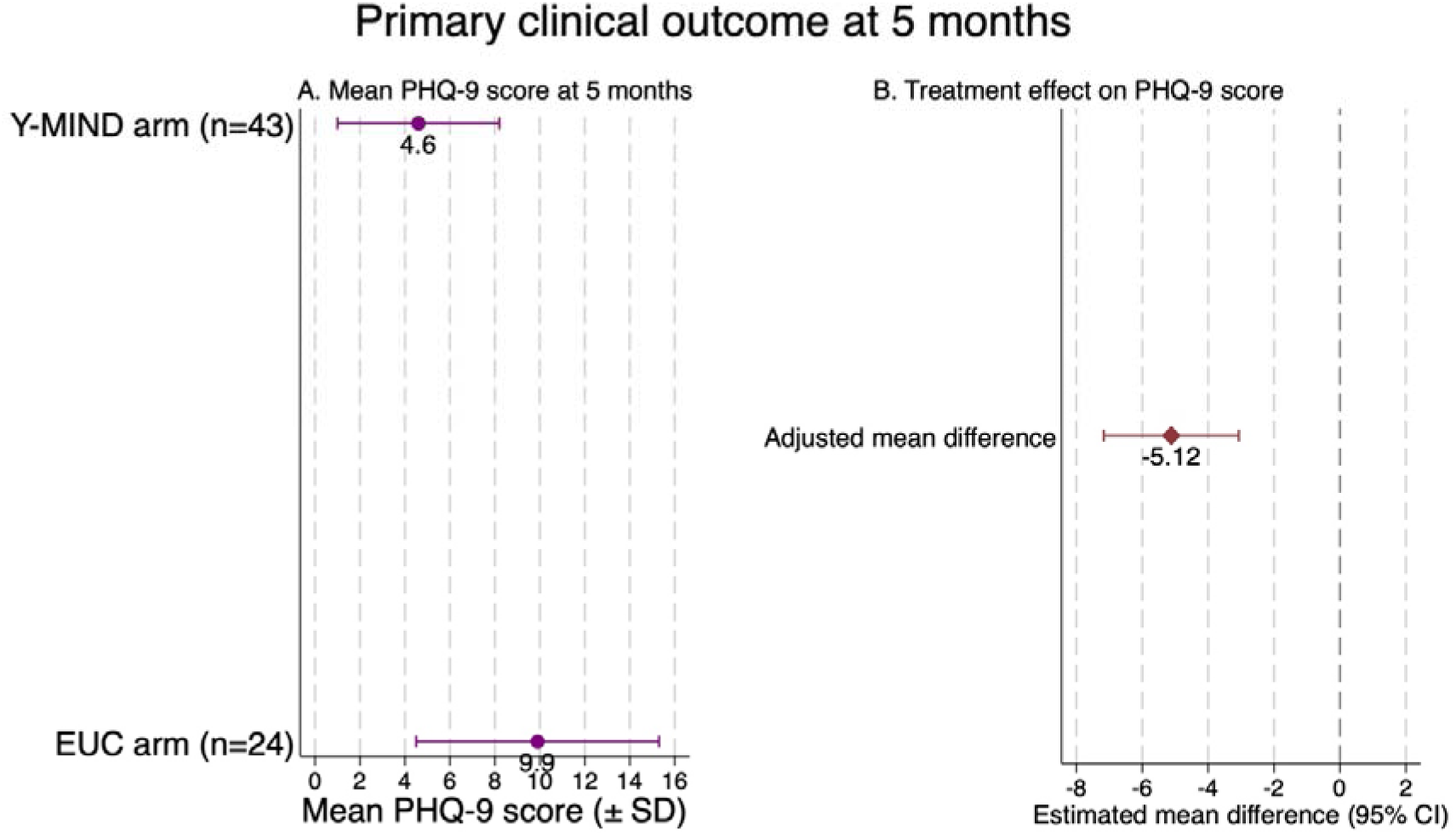
Effect of Y-MIND compared to EUC on primary and secondary clinical outcomes at 5 months.

The estimated first-year implementation cost was GHC 195,907 (USD 13,796), comprising start-up costs of GHC 106,307 (54%; USD 7,486) and recurrent costs of GHC 89,600 (46%; USD 6,310). Major start-up cost drivers were counselling room set-up (logistics and repairs) and training activities, which together accounted for over half of total implementation costs, while recurrent costs were dominated by counsellor responsibility allowances and printing of intervention materials. Personnel costs represented the largest single cost component, accounting for approximately 34% of total first-year expenditure.

**Table 2.** Summary of estimated first-year Y-MIND implementation costs across six senior high schools.

| Cost category | Cost (GHC) | Cost (USD) | % of total cost |
| --- | --- | --- | --- |
| Start-up costs | 106,307 | 7,486 | 54% |
| Recurrent costs | 89,600 | 6,310 | 46% |
| **Total first-year cost** | **195,907** | **13,796** | **100%** |

## Discussion

We report on the feasibility of Y-MIND, a brief psychological treatment delivered by Guidance and Counselling Coordinators (teachers) to adolescents in senior high schools with probable moderate to severe depression in Ghana. Overall, the findings indicate that both adolescents and GCCs rated Y-MIND as feasible, acceptable, and appropriate for delivery in real-world, low-resource school settings. We have four main findings.

Our first key finding is that trained Guidance and Counselling Coordinators (GCCs) were able to deliver Y-MIND by doing what matters in psychotherapy – “doing the right things well” (33). This was reflected not only in high fidelity and competence but also in strong implementation outcomes across feasibility, acceptability, and appropriateness. Both GCCs and adolescents perceived Y-MIND as relevant, usable, and well aligned with existing school-based roles and routines. Importantly, this was achieved with existing resources – teachers, reinforcing the potential for integrated care within existing education and health systems. What makes this finding particularly noteworthy is that prior research on task-shifting mental health care to teachers and similar non-specialist providers has documented significant implementation challenges in school settings, with teachers reporting limited preparation, competing role demands, and barriers to sustained delivery (34). Our qualitative evaluation (not reported here) identified time constraints as a potential barrier; however, this did not prevent the GCCs from delivering Y-MIND with high levels of competency and fidelity, though a similar trial in India showed the teacher-led arm encountered practical delivery constraints such as time and role-fit (35). We suggest that this was possible in Ghana because the GCCs already have a defined role of providing frontline psychosocial support, identifying students’ mental health needs, and facilitating referrals. Our programme strengthened the existing GCC role by equipping counsellors to deliver evidence-based psychological treatments in a structured and supervised manner, while creating an enabling environment through ongoing supervision and a dedicated counselling space. Supportive supervision is a critical component of lay-provider-delivered psychological interventions, as it promotes intervention fidelity and strengthens overall effectiveness (36). The usefulness of manualized psychological treatments in improving fidelity has been confirmed in previous research (37). Our study adds to the growing evidence supporting the involvement of teachers or teacher-like personnel in the provision of mental health interventions in low and middle-income countries in South-East Asia (India, Pakistan) (35,38,39), and Africa (Nigeria) (40).

Our second major finding is that it is feasible to conduct a fully powered effectiveness trial – achieved high consent and retention rates, successfully delivered all six Y-MIND sessions within the planned six-week window, and collected both clinical and self-report outcome data with minimal loss to follow-up. Importantly, these positive trial indicators may in part reflect our deliberate investment in rigorous eligibility verification prior to formal enrolment, which is consistent with established guidance on the role of feasibility studies in preparing for definitive trials (41,42).). We demonstrated the operational feasibility and efficiency of two-stage depression screening using the PHQ-2 as an initial screen followed by the PHQ-9 among those screening positive. Similar approaches have been evaluated in routine care settings in Uganda and Malawi (43,44) and recommended for adolescent screening to reduce respondent burden while retaining clinical detail (45). This improved recruitment efficiency. Of 290 adolescents initially screened, only 108 (37.2%) required administration of the full PHQ-9, reducing the number of full assessments by 62.8%. Among those screening positive on the PHQ-2, 67 (62.0%) met the trial eligibility criterion (PHQ-9 >9), demonstrating that the pre-screening approach effectively enhanced the pool of potentially eligible participants while reducing participant and researcher burden. Further, we employed structured eligibility adjudication techniques through a convening of a multidisciplinary eligibility review team (including investigators, country leads, project managers, and clinical psychologists). Whilst this added time to the recruitment process, it ensured that only participants who clearly met eligibility criteria were enrolled. Such procedures are often abbreviated in pragmatic settings due to efficiency and feasibility considerations; however, our favourable trial indicators are consistent with methodological literature suggesting that careful eligibility determination and participant selection may strengthen trial implementation, improve participant-intervention fit, and enhance the validity and reliability of trial findings (46).

Another key finding is the preliminary indication that Y-MIND can be implemented at a modest cost. Although a full economic evaluation was beyond the scope of this feasibility study, estimation of implementation costs aligns with recommendations for intervention development (47). From a public sector perspective, implementation costs were estimated at GHC195,907 ($13,796) per district and GHC32,651(USD2,299) per school. Given that Y-MIND is an intersectoral school-based intervention between the education and health sectors, implementation costs would be shared across sectors. In addition, integration of Y-MIND within existing systems suggests potential for scalability and sustainability with limited additional resource requirements, although further research is needed to confirm it.

Yet another encouraging finding is that, we observed initial clinical benefits, including lower severity of depressive symptoms among adolescents in the Y-MIND group as compared to those in the EUC group. The emergence of potential clinical benefit within a low-cost, non-specialist delivery model strengthens the case for Y-MIND as a scalable solution to adolescent depression in settings with limited specialist capacity. This finding is perhaps unsurprising given that Y-MIND was adapted from the original Youth Friendship Bench, which is grounded in problem-solving therapy (PST) – an evidence-based approach shown to reduce the severity of depression (28). The model is further strengthened by the incorporation of behavioural activation therapy, which in comparative adult studies has been associated with more than a 50% greater reduction in depressive symptom severity relative to prior medication-based treatment (48). Evidence for behavioural activation for adolescent depression in LMICs is emerging but not yet definitive (49).

### Strengths and limitations

This study has several important strengths. First, it represents one of the few cluster randomised feasibility trials evaluating a school based psychological intervention for adolescent depression in sub-Saharan Africa. The intervention was embedded within existing educational structures and delivered by Guidance and Counselling Coordinators who had no prior professional mental health training, thereby reflecting a realistic implementation pathway for scale up within routine systems. Second, the study demonstrated strong implementation outcomes, including high recruitment, complete acceptance among eligible participants, high retention at follow-up, and high treatment completion rates. Third, intervention fidelity and therapist competence were systematically assessed and demonstrated consistently high levels of adherence and quality of delivery. Fourth, the trial incorporated implementation outcomes including feasibility, acceptability, and appropriateness alongside clinical outcomes, which provided a thorough assessment of intervention readiness for a future definitive trial.

Several limitations should also be considered. As a feasibility study, the trial was not designed or powered to provide definitive evidence of effectiveness, and the observed clinical benefits should therefore be interpreted cautiously. The study included only six school clusters, which limited statistical precision and resulted in non-convergence of the prespecified mixed effects models, necessitating alternative analytical approaches. Although sensitivity analyses produced similar findings, estimates remain subject to uncertainty. Baseline imbalances were observed between trial arms, particularly in gender distribution and some socioeconomic characteristics. These differences likely arose from the small number of clusters and the cluster randomisation process rather than systematic recruitment bias, but they may have influenced outcome estimates. Finally, the rigorous eligibility adjudication procedures used in this feasibility trial enhanced diagnostic certainty and participant intervention fit but may be difficult to replicate in routine implementation settings without additional resources.

These limitations are typical of feasibility studies and do not undermine the primary objective of establishing the feasibility, acceptability, appropriateness, and implementation potential of Y-MIND prior to a fully powered effectiveness trial.

Some key lessons emerged with implications for the design of a future full trial. Whilst acknowledging the gender in-balance as a recruitment artifact, a future trial must ensure the selection of schools within the sampling frame accounts for a balance of single and mixed sex schools. We adhered to the statistical analysis plan as is expected; however, we encountered unexpected challenges with the data at analysis and had to contend with some deviations, which we carefully documented. Whilst it may be challenging to pre-empt such challenges, it may be useful in the planning of a future trial to build in strategies within the statistical analysis plan to accommodate such occurrences. Furthermore, piloting the trial procedures before the main trial proved useful. For example, we identified challenges in obtaining parental consent in a timely manner (particularly within boarding schools). In response, we strengthened the consent process by incorporating information sessions into parent–teacher meetings, which improved the consent rates.

## Conclusion

Taken together, these findings demonstrate that Y-MIND is feasible to deliver and is acceptable and appropriate for adolescents in Ghana, supporting progression to a fully powered effectiveness trial. While preliminary clinical findings are encouraging, this feasibility study was not designed to establish effectiveness. Future research should evaluate clinical and cost-effectiveness alongside implementation outcomes, including sustainable supervision models, and strategies to support long-term integration within Ghana’s school health system. Y-MIND could provide a scalable model for expanding access to evidence-based school mental health services in Ghana and other low- and middle-income countries.

## Data Availability

All data produced in the present study are available upon reasonable request to the authors

## AI Statement

The Authors declare that they have not used AI in any capacity that would require disclosure in accordance with Wiley’s AI Guidelines, nor in any capacity that would reasonably require discloser for editors, reviewers and readers to properly evaluate their research or manuscript. The authors confirm that AI has not been used for purposes including but NOT limited to drafting and editing, to generate substantial text or restructure arguments, nor in the research methodology. The authors take full responsibility for the accuracy of this statement.

## Acknowledgements

This research was funded by a National Institutes of Health Research grant to DC and MA (NIHR133384). We acknowledge the critical role of other members of the Y-MIND consortium (Young Person’s Advisory Group). We also acknowledge the generous partnership and support from the Ghana Education Service and the Ghana Health Service Directorates of the Navrongo Municipality and District.

