## supplementary table 1 for "A lay teacher-counsellor delivered brief psychological treatment for moderate to severe depression among older adolescents in senior high schools in Ghana (Y-MIND): a feasibility cluster randomized controlled trial": CONSORT-SPI-2018-Checklist.docx

| **SECTION** | **ITEM #** | **CONSORT 2010** | **CONSORT-SPI**  **2018** | **REPORTED ON PAGE #** |
| --- | --- | --- | --- | --- |
| **TITLE AND ABSTRACT** | | | | |
|  | 1a | Identification as a randomised trial in the title^§^ |  | 1 |
|  | 1b | Structured summary of trial design, methods, results, and conclusions (for specific guidance see CONSORT for Abstracts)^§^ | Refer to CONSORT extension for social and psychological intervention trial abstracts | 3 |
| **INTRODUCTION** | | | | |
| Background and  Objectives | 2a | Scientific background and explanation of rationale ^§^ |  | Introduction, page 4-5 |
|  | 2b | Specific objectives or hypotheses ^§^ | If pre-specified, how the intervention was hypothesied to work | Introduction, page 5 |
| **METHODS** | | | | |
| Trial Design | 3a | Describe of trial design (such as parallel, factorial), including allocation ratio ^§^ | If the unit of random assignment is not the individual, please refer to CONSORT for Cluster Randomized Trials | Methods, page 7 |
|  | 3b | Important changes to methods after trial commencement (such as eligibility criteria), with reasons |  | N/A |
| Participants | 4a | Eligibility criteria for participants^§^ | When applicable, eligibility criteria for settings and those delivering the interventions | Methods, page 6 |
|  | 4b | Settings and locations where the data were collected |  | Methods, page 6 |
| Interventions | 5 | The interventions for each group with sufficient details to allow replication, including how and when they are actually administered ^§^ |  | Methods, page 9 |
|  | 5a |  | Extent to which interventions were actually delivered by providers and taken up by participants as planned | Methods, page 10 |
|  | 5b |  | Where other informational materials about delivering the intervention can be accessed | Protocol paper, reference list number 7 |
|  | 5c |  | When applicable, how intervention providers were assigned to each group | N/A |
| Outcomes | 6a | Completely defined pre-specified outcomes, including how and when they were assessed^§^ |  | Methods, page 10 |
|  | 6b | Any changes to trial outcomes after the trial commenced, with reasons |  | N/A |
| Sample Size | 7a | How sample size was determined^§^ |  | Methods, page 7 |
|  | 7b | When applicable, explanation of any interim analyses and stopping guidelines |  | N/A |
| **RANDOMISATION** | | | | |
| Sequence  generation | 8a | Method used to generate the random allocation sequence |  | Methods, page 7 |
|  | 8b | Type of randomisation; detail of any restriction (such as blocking and block size)^§^ |  | Methods, page 7 |
| Allocation concealment mechanism | 9 | Mechanism used to implement the random allocation sequence, describing any steps taken to conceal the sequence until interventions were assigned^§^ |  | Methods, page 7 |
| Implementation | 10 | Who generated the random allocation sequence, who enrolled participants, and who assigned participants to interventions^§^ |  | Methods, page 7 |
| Awareness of assignment | 11a | Who was aware of intervention assignment after allocation (for example, participants, providers, those assessing outcomes), and how any masking was done |  | Methods, page 7,11 |
|  | 11b | If relevant, description of the similarity of interventions |  | Methods, page 8,9 |
| Analytical  methods | 12a | Statistical methods used to compare group outcomes^§^ | How missing data were handled, with details of any imputation method | Methods, page 12 |
|  | 12b | Methods for additional analyses, such as subgroup analyses, adjusted analyses, and process evaluations |  | Methods, page 12,13 |
| **RESULTS** | | | | |
| Participant flow (a diagram is strongly recommended) | 13a | For each group, the numbers randomly assigned, receiving the intended intervention, and analysed for the outcomes^§^ | Where possible, the number approached, screened, and eligible prior to random assignment, with reasons for non-enrolment | Results Fig 1 |
|  | 13b | For each group, losses and exclusions after randomisation, together with reasons^§^ |  | Results Fig 1 |
| Recruitment | 14a | Dates defining the periods of recruitment and follow-up |  | Methods 7 |
|  | 14b | Why the trial ended or was stopped |  | N/A |
| Baseline data | 15 | A table showing baseline characteristics for each group^§^ | Include socioeconomic variables where applicable | Results, page 14 |
| Numbers analysed | 16 | For each group, number included in each analysis and whether the analysis was by original assigned groups^§^ |  | Results Fig 1 |
| Outcomes and estimation | 17a | For each outcome, results for each group, and the estimated effect size and its precision (such as 95% confidence interval)^§^ | Indicate availability of trial data | Results Page 14-16 |
|  | 17b | For binary outcomes, the presentation of both absolute and relative effect sizes is recommended |  | Results Page 14-16 |
| Ancillary analyses | 18 | Results of any other analyses performed, including subgroup analyses, adjusted analyses, and process evaluations, distinguishing pre-specified from exploratory |  | Results Page 16 |
| Harms | 19 | All important harms or unintended effects in each group (for specific guidance see CONSORT for Harms) |  | N/A |
| **DISCUSSION** | | | | |
| Limitations | 20 | Summarize the main results (including an overview of concepts, themes, and types of evidence available), link to the review questions and objectives, and consider the relevance to key groups. | Trial limitations, addressing sources of potential bias, imprecision, and, if relevant, multiplicity of analyses | Discussion 17-20 |
| Generalisability | 21 | Discuss the limitations of the scoping review process. | Generalisability (external validity, applicability) of the trial findings^§^ | Discussion 20 |
| Interpretation | 22 | Provide a general interpretation of the results with respect to the review questions and objectives, as well as potential implications and/or next steps. | Interpretation consistent with results, balancing benefits and harms, and considering other relevant evidence | Discussion 17-20 |
| **IMPORTANT INFORMATION** | | | | |
| Registration | 23 | Registration number and name of trial registry |  | Abstract, page 4 |
| Protocol | 24 | Where the full trial protocol can be accessed, if available |  | Reference List 7 |
| Declaration of Interests | 25 | Sources of funding and other support; role of funders | Declaration of any other potential interests | Acknowledgement, page 22 |
| Stakeholder investments | 26a |  | Any involvement of the intervention developer in the design, conduct, analysis, or reporting of the trial | Yes |
|  | 26b |  | Other stakeholder involvement in trial design, conduct, or analyses | Yes |
|  | 26c |  | Incentives offered as part of the trial | Yes |

This table lists items from the CONSORT 2010 checklist (with some modifications for social and psychological intervention trials) and additional items in the CONSORT-SPI 2018 extension. Empty rows in the ‘CONSORT-SPI 2018’ column indicate that there is no extension to the CONSORT 2010 item

*We strongly recommended that the CONSORT-SPI 2018 Explanation and Elaboration (E&E) document be reviewed when using the CONSORT-SPI 2018 checklist for important clarifications on each item

§An extension item for cluster trials exists for this CONSORT 2010 item

This checklist is derived from:

- Montgomery, P., Grant, S., Mayo-Wilson, E., Macdonald, G., Michie, S., Hopewell, S., & Moher, D. (2018). Reporting randomised trials of social and psychological interventions: the CONSORT-SPI 2018 Extension. *Trials*, *19*(1), 407.
- Grant, S., Mayo-Wilson, E., Montgomery, P., Macdonald, G., Michie, S., Hopewell, S., & Moher, D. (2018). CONSORT-SPI 2018 Explanation and Elaboration: guidance for reporting social and psychological intervention trials. *Trials*, *19*(1), 406.
- Schulz, K. F., Altman, D. G., & Moher, D. (2010). CONSORT 2010 Statement: updated guidelines for reporting parallel group randomised trials. *BMJ*, *340*, c332.

Montgomery 2018 and Grant 2018 were distributed under the terms of the Creative Commons Attribution 4.0 International License ([http://creativecommons.org/licenses/by/4.0/](https://urldefense.com/v3/__http:/creativecommons.org/licenses/by/4.0/__;!!C5qS4YX3!CdcaroZO5hxp8FYacDXbUMpysBqFhPEDSGHr-PBP-6JeLAF0ufQ5lz_kopWiil164numaGA4JU3gaxY-WqOg1G1v39F-mLM$)). Schulz 2010 was distributed under the terms of a Creative Commons Attribution Non-commercial License (<https://creativecommons.org/licenses/by-nc/2.0/>). We have revised the checklists as published to include an extra column for “reporting on page #”.
